# COGNET-STROKE: A NOVEL BRAIN DYSCONNECTIVITY TOOL FOR PREDICTION OF COGNITIVE DEFICITS AFTER STROKE

**DOI:** 10.1101/2025.08.13.25333589

**Authors:** Antonio Jimenez-Marin, Silke Boulanger, Iñigo Tellaetxe-Elorriaga, Iñaki Escudero, Alberto Cabrera-Zubizarreta, Gil De Sousa, Marimar Freijo, Pedro I. Tejada, Roberto Toro, Ibai Diez, Asier Erramuzpe, Jesus M. Cortes

## Abstract

This study presents COGNET-STROKE, a novel meta-analytic brain functional decoding tool designed to predict cognitive deficits after stroke. The tool integrates Lesion Network Mapping with meta-analytic concept maps derived from Neurosynth, enabling the identification of the cognitive domains most affected by stroke-induced network disruptions. Validation analyses confirmed that identified patients with predicted motor and sensory impairment had significantly higher scores in their corresponding NIHSS-subscales, demonstrating predictability from lesion-induced dysconnectivity to behavioral impairment. COGNET-STROKE openly available at https://github.com/compneurobilbao/COGNET-STROKE, offers a framework for individualized cognitive-deficit profiling, with implications for personalized rehabilitation strategies in stroke recovery.

## 1 Introduction

Stroke defined as, *“rapidly developing clinical signs of focal (or global) disturbance of cerebral function, with symptoms lasting 24 hours or longer, or leading to death, with no apparent cause other than of vascular origin”* (Aho et al., 1980), is a leading cause of death and disability worldwide. With an annual mortality rate of about 5.5 million and up to 50 % of survivors experiencing chronic disability, stroke has a profound impact on public health, carrying significant social and economic consequences (Caplan, 2016; Lopez et al., 2006; Warlow, 1998). This burden is expected to rise substantially as an effect of ongoing demographic changes; like ageing of the population and health transitions (Amuna & Zotor, 2008; Bosu, 2010; Donkor, 2018; Owolabi et al., 2015). Moreover, therapeutic advances have increased stroke survival rates, particularly in developed countries, highlighting the urgent need for enhanced prevention, treatment, and rehabilitation strategies (A. S. Kim et al., 2015; Pu et al., 2023). A critical aspect of optimizing stroke care lies in understanding how brain lesions impact behavior. Clinical decision-making in stroke management depends on accurately linking stroke-induced lesions to resulting functional impairments. Traditionally, Lesion Symptom Mapping (LSM) has been employed to associate specific symptoms with corresponding brain lesions (Broca, 1861; Moore & Demeyere, 2020; Ranzini et al., 2023; Wernicke, 1974). However, inter-individual variability in lesion size and location hinders precise lesion-symptom mapping, underscoring the need for more granular, network-based approaches to optimally guide targeted interventions. Increasing evidence supporting the connectome hypothesis suggests that symptoms arise not from focal damage alone but from disruptions in specific brain networks. This shift has fueled the rise of lesion-driven dysconnectivity analyses and the development of Lesion Network Mapping (LNM) (Boes et al., 2015; M. D. Fox, 2018), a standardized computational framework that connects lesion-induced disconnection to behavioral outcomes. The LNM framework allows for insights on the brain networks that are disconnected by the lesion, and has provided valuable insights into behavioral symptoms that were previously challenging to localize with other methods such as tremor (Joutsa et al., 2019) or hallucinations (N. Y. Kim et al., 2021). Furthermore, LNM has recently proved successful in predicting stimulation sites to improve symptoms, such as addiction (Padmanabhan et al., 2019) or tics (Ganos et al., 2022). Recent work has highlighted that when atrophy sites or lesions are highly spatially heterogeneous, the common LNM estimated across patients can become strongly constrained by intrinsic topological properties of the reference connectome. In particular, averaging across many distinct lesions tends to produce population-level disconnection maps that progressively converge toward the brain’s degree distribution, emphasizing highly connected hub regions rather than symptom-specific network alterations (Van Den Heuvel et al., 2026). Nevertheless, studies incorporating complementary sensitivity and specificity analyses have demonstrated that, beyond these degree-related contributions, normative disconnection patterns retain substantial symptom-specific information independent of degree maps (Meng et al., 2026; Siddiqi et al., 2026). Thus, although part of the association with clinical symptoms may reflect a common contribution from high-degree hub regions, higher-order network effects preserve sufficient amount of specificity with respect to the symptom or behavioral phenotype being modeled. On the other hand, LNM studies have traditionally relied on task performance or symptom linking lesion network disruptions to their associated deficits (Joutsa et al., 2022). This approach presents the major challenge of causal inference. For example, if a brain region is broadly activated across multiple tasks, its activation alone offers limited insight into the specific cognitive processes that may be disrupted in patients. As a result, attributing a particular symptom to that brain region may be misleading. This challenge, known as the *reverse inference problem*, highlights the difficulty of deducing a person’s mental state solely from patterns of brain activation (Poldrack, 2011). To address this problem, researchers have developed large-scale meta-analytic databases of fMRI studies (Cortese et al., 2016; Rubia, 2016), such as BrainMap (P. T. Fox & Lancaster, 2002), Neurosynth (Yarkoni et al., 2011), and NeuroQuery (Dockès et al., 2020). These databases enable functional decoding of specific brain regions, offering a data-driven approach to linking brain activation with mental processes (Reetz et al., 2012). Instead of relying solely on direct symptom mapping, functional decoding could allow for a more systematic interpretation of lesion-induced disruptions in brain networks. In this study, we present COGNET-STROKE, combining precise dysconnectivity maps obtained by LNM with meta-analytic functional decoding obtained by Neurosynth to generate cognitive concept-related brain maps which are most affected by lesion-induced network disruptions. By linking them to broader cognitive functions, rather than isolated symptoms, COGNET-STROKE provides a more comprehensive, data-driven framework for understanding stroke-related cognitive deficits. Ultimately, we present a validation for COGNET-STROKE limited to two domains, i.e. motor and sensory, verifying that predicted impairment corresponds to behavioral deficits for motor and sensorial domains, and further research is needed for other cognitive and behavioral domains. COGNET-STROKE is useful for clinicians to facilitate individualized rehabilitation strategies, allowing for tailored cognitive rehabilitation interventions based on an individualized patient-specific network disruption.

## 2 Methods

### 2.1 Participants

This study was approved by the Clinical Research Ethics Committee of Cruces University Hospital (code E19/52, PI: Jesus M Cortes). This is a retrospective study involving patients with stroke aiming to associate brain imaging to rehabilitation outcome. Eligible participants included all patients admitted to the Gorliz Hospital during the chronic-phase of stroke who underwent a rehabilitation program. A total of 290 hemorrhagic stroke patients were included for the study, with a mean age of 70.5 years (*σ* = 13.2, range 19 – 97), including 169 males.

### 2.2 Assessments and behavioral scores

Functional assessments pre and post rehabilitation were conducted by Hospital Staff using six standardized clinical tests:

1. Barthel Index: A measure of functional independence that evaluates the ability to perform 10 activities of daily living (ADLs), including feeding, bathing, dressing, and mobility (Mahoney & Barthel, 2013). Scores range from 0 (fully dependent on others for performing ADLs) to 100 (no significant disabilities in basic self-care functions).
2. National Institutes of Health Stroke Scale (NIHSS): A standardized neurological assessment tool that quantifies stroke severity based on motor function, sensory deficits, speech, and level of consciousness (Brott et al., 2016). NIHSS scores range from 0 (no neurological deficits) to 42 (severe signs of stroke-related impairments in the assessed functions).
3. Trunk Control Test (TCT): A rehabilitation test measuring four aspects of trunk movement; rolling to the weak and strong side, balance in sitting position and sitting up from lying down. Scores range from 0 (no trunk control) to 100 (no impairments concerning trunk control) (Collin & Wade, 1990).
4. Functional Oral Intake Scale (FOIS): An ordinal scale assessing the extent to which a patient can safely consume food and liquids by mouth (Crary et al., 2005). FOIS is a 7-point scale ranging from 1 (the patient is not able to do any oral intake) to 7 (a total oral diet with no restrictions is possible).
5. Pfeiffer Test: A cognitive screening tool designed to detect cognitive impairment or early signs of dementia (Pfeiffer, 1975). Every incorrect answer on the test adds one point, with 0 points being normal cognitive function, and maximum 10 points indicating severe cognitive impairment.
6. Functional Ambulation Categories (FAC): A five-point scale used to assess a patient’s level of independent walking ability. A score of 0 indicates a complete inability to walk, while a score of 5 reflects full ambulation without assistance.

### 2.3 Image acquisition

All the patients had a computed tomography (CT) acquired at the acute-phase hospital admission. For the generation of LNM-derived functional connectivity maps, images from healthy subjects were obtained from the Human Connectome Project Dataset (HCP) (Van Essen et al., 2012). This dataset is recognized for its large number of participants (N ∼ 1200), high-quality multimodal acquisitions and includes preprocessed MRI sequences, making it well-suited for connectivity analyses.

### 2.4 Lesion segmentation and coregistration

Lesions were manually segmented using 3D Slicer software (version 5.6.2; https://www.slicer.org/). All segemented lesions were supervised by an experienced neuroradiologist (IE). After that, the lesions masks were validated by an independent experienced neuroradiologist (ACZ). To register the masks to the MNI152 template, we developed a tool (https://github.com/compneurobilbao/CTLesion2MNI152) based on (Kuijf et al., 2013; Po-Yu Kao, 2019). Very briefly, our tool detects the skull on the CT based on the Hounsfield Units (HU *>* 100) and computes an affine registration matrix to the MNI152 skull. Then the tool extracts the brain (HU *>* 0, HU *<* 100) and applies the affine registration, with the addition of a nonlinear warp calculated without consideration of the lesion area for avoiding lesion-derived distortions that can worsen the registration quality.

### 2.5 Construction of Lesion-Induced Functional Dysconnectivity Maps

To successfully map stroke lesions to their respective dysconnectivity patterns, resting-state functional MRI (rs-fMRI) data from healthy controls aged 22 to 37 (n = 1000) were obtained from the HCP dataset. The data was preprocessed with ICA-FIX (Salimi-Khorshidi et al., 2014), and additionally we applied Global signal regression, band-pass filtering between 0.01-0.08Hz and spatial smoothing with FWHM of 6mm. The functional connectivity maps for each stroke patient were generated through seed-based connectivity (SBC) analysis following the pipeline described in (Jimenez-Marin et al., 2022). Summarizing, the lesion mask of each stroke patient registered to the MNI152 template was used as the seed region for SBC analyses, which were conducted separately for each HCP subject. Pearson correlation coefficients (r) were computed between the mean timeseries of the seed region —obtained by averaging voxel-wise time-series within each lesion— and the time-series of all other brain voxels. To improve normality, these r-values were Fisher-transformed using the inverse hyperbolic tangent function. This process resulted in a 3D brain map of z-values for each stroke lesion, generated independently for each HCP subject. A one-sample t-test was then applied across the 1000 individual connectivity maps to derive a final functional disconnection map for each lesion, yielding a group-level statistical representation of lesion-induced functional dysconnectivity. The pipeline is openly available at https://github.com/compneurobilbao/lnm. These LNM-derived functional dysconnectivity maps contain both positive and negative z-values, each carrying distinct theoretical interpretations. Positive z-values indicate areas that generally exhibit correlated activity with the lesioned region, while negative z-values represent areas where activity is anticorrelated with the lesioned region. Given these contrasting implications, each patient’s functional dysconnectivity maps were divided into two separate representations: one for lesion-correlated connectivity (positive values) and one for lesion-anticorrelated connectivity (negative values).

### 2.6 Derivation of meta-analytic cognitive-concept-related brain maps

To generate cognitive-concept-related brain maps, we made use of the Neurosynth database (Yarkoni et al., 2011), which employs an automated text-mining approach to extract activation coordinates from published neuroimaging studies. Neurosynth processes the full text of each study to identify frequently occurring terms, which are then assigned to the respective study. Only terms appearing in at least 20 studies are included in the database. Subsequently, every term included in the Neurosynth database can be used to create a term-specific brain map, with values reflecting the likelihood of activation in a specific brain area given that term (see Fig 1).

**Figure 1:**
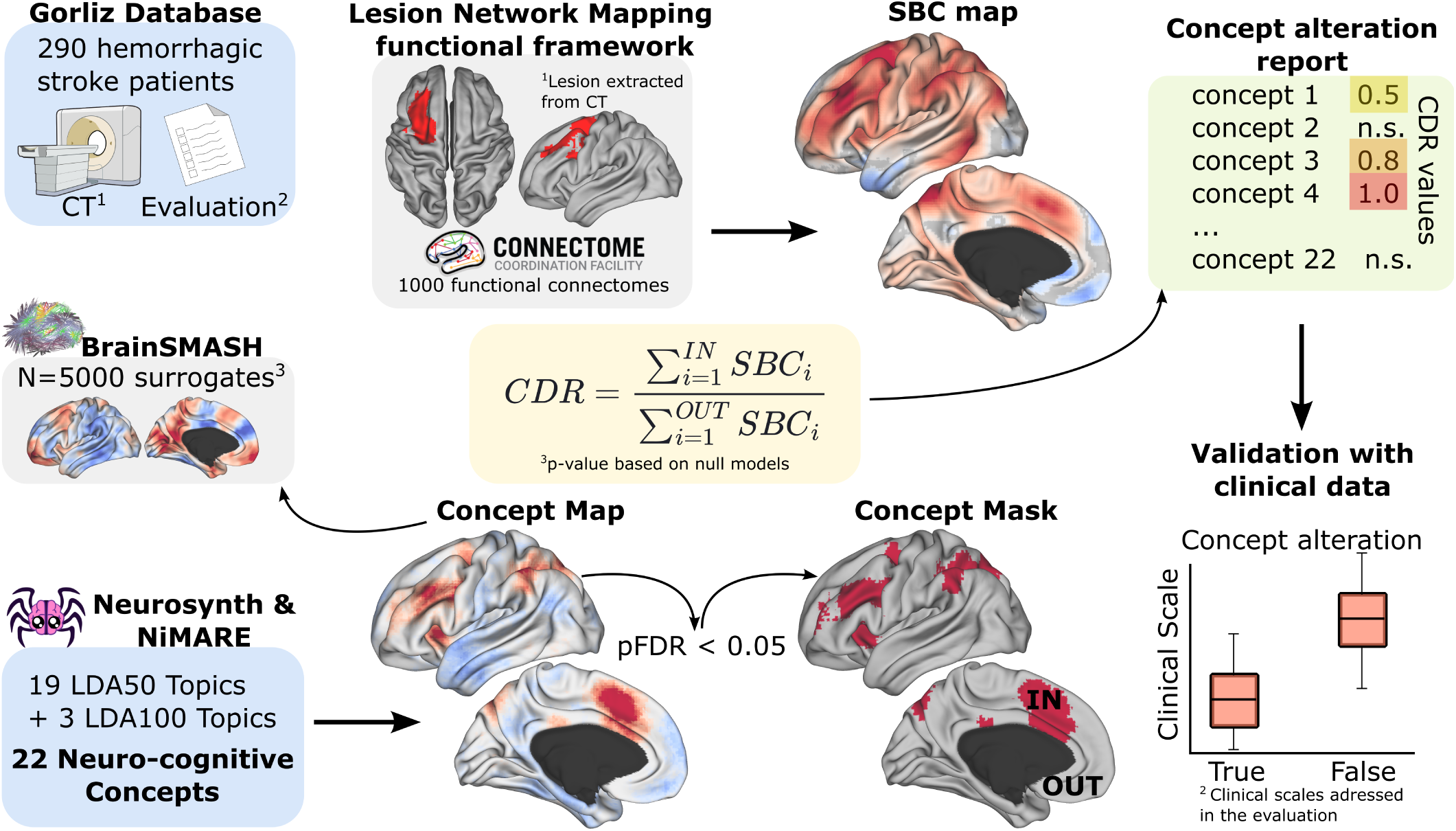
Pipeline followed by COGNET-STROKE, to obtain and validate the concepts affected of a stroke participant. Lesion network mapping was applied to obtain individual seed based correlation (SBC) maps. Together with the concept maps obtained from the Neurosynth database using the NiMARE software package, we compute the concept to dysconnectivity representation (CDR) values. We used the BrainSMASH tool to generate null-maps in order to calculate the significance of the CDR value. Finally, using the available clinical evaluation of the patients, we validate the CDR obtained as the comparison of the clinical score between patients with significant CDR and patients without it.

However, using the standard Neurosynth database presents two challenges: the sheer number of terms and the presence of non-cognitive terms such as “gyrus,” “woman,” or “cortex”. To address the first problem, we followed the approach of (Margulies et al., 2016) and derived our overarching concept-related brain maps from the 50 set of topic bins (v5) or 100 set of topic bins (v5) provided by Neurosynth’s creators. These bins were created by applying Latent Dirichlet Allocation (LDA) to group all Neurosynth database terms into topic bins (Poldrack et al., 2012). This approach ensured that our concepts were comprehensive and representative of broader cognitive constructs, rather than being limited to single-term associations. By leveraging topic bins, we preserved the richness of conceptual information while minimizing the risk of data loss due to overly restrictive filtering. For example, if we were to relate our concept brain map to only a single Neurosynth database term, such as “attention,” we might inadvertently exclude studies that use related but distinct terms like “focus,” “vigilance,” or “concentration.” By using topic bins, we ensured that our concept maps captured a richer and more comprehensive representation of cognitive processes, reducing the risk of missing relevant data and improving the robustness of our analyses. The second challenge was addressed by determining which terms are cognitively relevant. We analyzed the 50 term-based bins and identified 19 as cognitively meaningful. This identification was based on the neuro-behavioral concepts listed in (Luppi et al., 2022; Margulies et al., 2016), which were updated to match the Neurosynth database version 5 state. Additionally, to enhance the granularity of the concepts ‘auditory perception’, ‘multisensory integration’ and ‘tactile perception, we used the more detailed 100-bin model. In total, this process resulted in 22 cognitively relevant concepts and corresponding concept maps for analysis. For a detailed description and definition of all the concepts and their origin, refer to Table 1.

**Table 1:** Definitions, origin and top LDA weighted terms of each concept.

| Concept | Topic number bins | Top 3 LDA weights | Definition |
| --- | --- | --- | --- |
| Motor | 17 (50 bins) | Motor, cortex, hand | "The function of supervising motor activities" (Poldrack et al., 2011) |
| Eye movements | 44 (50 bins) | Eye, sleep, gaze | "Any shift of position of the eye in its orbit" (Sereno & Bolding, 2017) |
| Motion Perception | 45 (50 bins) | Motion, perception, visual | "The ability to perceive and interpret the movement of objects, including self-motion, as well as the guidance of eye and hand movements" (Barton, 2021) |
| Auditory perception | 81 (100 bins) | Auditory, sounds, sound | "The ability to identify, interpret, and attach meaning to sound" (Poldrack et al., 2011) |

Table 1 continued
| Concept | Topic number<br>bins | Top 3<br>LDA<br>weights | Definition |
| --- | --- | --- | --- |
| Visual per-<br>ception | 42 (50 bins) | Visual, cortex,<br>sensory | "The ability to interpret the surrounding environment by processing information that is contained in visible light. The resulting perception is also known as eyesight, sight, or vision" (Poldrack et al., 2011) |
| Tactile per-<br>ception | 82 (100 bins) | Somatosensory,<br>stimulation,<br>tactile | "The perception of the qualities and properties of material surfaces by touch" (Okamoto et al., 2013) |
| Pain | 32 (50 bins) | Pain, so-<br>matosensory,<br>stimulation | "An unpleasant sensory and emotional experience associated with actual or potential tissue damage" (Poldrack et al., 2011) |
| Action | 19 (50 bins) | Action, actions,<br>observation | "The bringing about of an alteration by force or through a natural agency; expression by means of attitude, voice, and gesture; a function of the body or one of its parts; an act of will; a thing done; the accomplishment of a thing usually over a period of time, in stages, or with the possibility of repetition" (Poldrack et al., 2011) |
| Facial per-<br>ception | 40 (50 bins) | Face, faces, fa-<br>cial | "The process by which faces are identified as such" (Poldrack et al., 2011) |
| Multisensory<br>integration | 97 (100 bins) | Visual, motion,<br>auditory | "A process by which information from different sensory systems is combined to influence perception, decisions, and overt behavior" (Stein et al., 2009) |
| Attention | 47 (50 bins) | Attention,<br>attentional,<br>target | "Used to describe any number of cognitive processes, organized as top-down or bottom-up, goal-directed or stimulus-driven, and more, but generally reflecting an interplay between cognitive and sensory systems" (Poldrack et al., 2011) |
| Working<br>memory | 9 (50 bins) | Memory, work-<br>ing, wm | "Active maintenance and flexible updating of goal/task relevant information (items, goals, strategies, etc.) in a form that resists interference but has limited capacity. These representations may involve flexible binding of representations, may be characterized by the absence of external support for the internally maintained representations, and are frequently temporary due to ongoing interference" (Barton, 2021) |
| Inhibition | 16 (50 bins) | Response, inhi-<br>bition, control | "The process by which a response or thought is suppressed" (Poldrack et al., 2011) |

Table 1 continued
| Concept | Topic number bins | Top 3 LDA weights | Definition |
| --- | --- | --- | --- |
| Memory | 33 (50 bins) | Memory, re-trieval, encoding | "The ability of an organism to use past events to inform/influence current actions" (Poldrack et al., 2011) |
| Language | 37 (50 bins) | Language, reading, word | "The mental ability to encode and decode information, and translate this information into verbal, acoustic and visual representations, according to a set of rules that are common across a population" (Poldrack et al., 2011) |
| Numerical cognition | 18 (50 bins) | Number, ips, numerical | "Set of cognitive processes pertaining to the assimilation, ascription and manipulation of numerical information" (Poldrack et al., 2011) |
| Cognitive control | 20 (50 bins) | Control, conflict, task | "The top-down modulation of cognitive processes based on higher-order representations such as goals or plans" (Poldrack et al., 2011) |
| Imagery | 41 (50 bins) | Imagery, mental, events | "A memory system consisting of episodes recollected from an individuals life, based on a combination of episodic (personal experiences and specific objects, people and events experienced at particular time and place) and semantic (general knowledge and facts about the world) memory" (Poldrack et al., 2011) |
| Social cognition | 28 (50 bins) | Social, empathy, moral | "The encoding, storage, retrieval, and processing, in the brain, of information relating to conspecifics, or members of the same species" (Poldrack et al., 2011) |
| Emotion | 26 (50 bins) | Emotional, amygdala, negative | "A complex of psychological phenomena that involve some degree of arousal and valence (positive/negative)" (Poldrack et al., 2011) |
| Decision making | 30 (50 bins) | Decision, making, risk | "The deliberate selection of a single alternative amongst multiple alternatives" (Poldrack et al., 2011) |
| Reward processing | 7 (50 bins) | Reward, feedback, striatum | "A positive return for performance of a specific behavior" (Poldrack et al., 2011) |

*Note.* All concepts are based on the concept used in the Neurosynth meta-analytic gradient analyses from (Luppi et al., 2022; Margulies et al., 2016).

Each of the bins, encompassing all associated terms, was used to query the Neurosynth database to create a single overarching concept map encompassing all the terms. For each bin, studies were categorized into two groups: those that contain the terms in the bin and those that do not. A large-scale multi-level kernel density (MKDA) meta-analysis was then performed, comparing reported activation coordinates between these groups to generate a z-statistic map for each cognitive term, reflecting the likelihood of activation in specific brain regions associated with that term. To correct for multiple comparisons, a false discovery rate (FDR) of 0.01 was used.

### 2.7 Resting-state network correspondece to meta-analytic cognitive-concept-related brain maps

As an additional control for the theoretical validity of our concepts, and to see whether correlating the concept maps to functional dysconnectivity maps was a valid approach, we calculated similar to (Gillig et al., 2025) the associations between our concepts with Resting State Network (RSN) data. In particular, we calculated correlations between the concept maps and different RSN maps using Neuromaps (Markello et al., 2022). To evaluate the significance of the correlations, we generate 5,000 null-maps of the concept maps using BrainSmash (Burt et al., 2020). To maximize the computational efficiency of the null-map generation process, all brain maps were parcellated into a whole-brain atlas with 2,507 regions of interest (ROIs), obtained from (Jimenez-Marin et al., 2024) and adding the cerebellum and brain-stem following their same methodology. The RSN maps were derived following (Tahedl & Schwarzbach, 2023), adding the dorsal attention, salience and language networks using the masks included in CONN toolbox (https://web.conn-toolbox.org/).

### 2.8 Concept to disconnection representation index

To link the dysconnectivity maps to each concept, we first split the dysconnectivity map in two, positive and negative dysconnectivity, corresponding respectively to areas with dynamics correlated or anticorrelated with the lesion dynamics. Similar to the procedure explained in the previous section, we parcellated the brain maps with 2,507 regions of interest (ROIs) and generated 5,000 null-maps for every concept map. Then, we generated a concept mask including all the voxels with pFDR *<* 0.05 in the map. We computed the concept to disconnection representation index (CDR) as the average of the dysconnectivity values inside each concept mask divided by the average of the dysconnectivity values outside each concept mask, separately for positive and negative dysconnectivities. Therefore, the measure do not depend on concept-mask size, broad dysconnectivity, or overall lesion burden. The CDR expresses how much a dysconnectivity pattern matches a given concept mask, assessing the potential alteration of cognitive functions related to the concept. Finally, we obtained a p-value per CDR using the null-maps, marking for each patient all the concepts with *p* − *value_bonferroni_ <* 0.05 as potentially affected.

### 2.9 Validation analysis

To test the clinical significance of model predictions, we examined the relationships between CDR and the clinical scores. From the NIHSS score, we extracted the score for motor and sensory performance. Motor score was the sum of motor arm and motor leg assessments with possible values 0-16, while sensory score was the sum of sensation subitem (tactile perception) plus extinction and inattention subitem (visual/tactile/auditory/spatial/personal innatention), with possible values 0-4. We computed the Wilcoxon rank-sum statistic (a non-parametric test for finding differences in two groups) in each sub-score between the group with CDR alteration (*p* − *value_bonferroni_ <* 0.05) and the one without alteration. We also compared the behaviour predictability from dysconnectivity maps with predictions based on the lesion location. Following a procedure similar to the one for obtaining the CDR, we compute an overlap between the concept mask and the lesion mask, using null-maps for obtaining the statistical significance of the overlap.

## 3 Results

This study aimed to create a tool to predict cognitive deficits from dysconnectivity maps after stroke. To do so, we quantitatively analyze the relationship between meta-analytically derived cognitive concept maps using Neurosynth and functional dysconnectivity maps generated generated by LNM. Twenty-two cognitively relevant concept-maps were defined based on (Margulies et al., 2016), and validated by examining their correlations to existing and theoretically well-known RSNs. Afterwards, we analyzed a cohort of 290 stroke patients, deriving 44 concept scores per patient, 22 from lesion-correlated and 22 from lesion-anticorrelated connectivity maps. The statisticalsignificance was assessed by a comparison to a brain-surrogate map null distribution. For validation, we compared behavioral scores between two patient groups: individuals for whom COGNET-STROKE predicted significant motor and sensory concept alterations and those for whom it predicted none. Fig 1 shows the pipeline followed to obtain and validate the concept to disconnection representations.

### 3.1 RSN-concept association

First, we studied the association between classical RSNs (see Methods) and cognitive concept maps (Fig 2). A diagonal trend was observed, where higher-order RSNs (left side) correlated more strongly with higher-order cognitive concepts (bottom), while lower-order RSNs (right side) showed stronger associations with sensory and motor-related concepts (top). The executive control network showed correlations to the concepts working memory (r = .29, p *<* .01), inhibition (r = .37, p *<* .001) and cognitive control (r = .36, p *<* .001). The concepts cognitive control (r = .33, p *<* .001), numerical cognition (r = 0.52, p *<* .001), language (r = .42, p *<* .001) and working memory (r =.50, p *<* .001) were correlated with the left frontoparietal network. The default mode network had a correlation to the concepts of imagery (r = .24, p *<* .001), memory (r = .34, p *<* .001) and social cognition (r = .31, p *<* .001). The language network had a significant correlation to the concept social cognition (r = .35, p *<* .001) and language (r = .48, p *<* .001). The salience network had correlations with concepts cognitive control (r = .35, p *<* .001), inhibition (r = 0.37, p *<* .001) and working memory (r =.50, p *<* .001). The dorsal attention network and the concepts motor (r = .30, p *<* .001), tactile perception (r = .37, p *<* .001), and action (r = .30, p *<* .001) were correlated. The lateral visual network had correlations to motion perception (r = .57, p *<* .001), to visual perception (r = .57, p *<* .001), to action (r = .39, p *<* .001), to multi-sensory integration (r = .46, p *<* .001), to attention (r = .55, p *<* .001), and to numerical cognition (r = .39, p *<* .001). The auditory network had correlations to auditory perception (r = .78, p *<* .001) to pain and multi-sensory integration (r = .33, p *<* .01). The occipital pole network only had correlation to motion perception (r = .28, p *<* .01) and visual perception (r = .33, p *<* .001). The sensory-motor network had correlations to motor (r = .44, p *<* .001) and tactile perception (r = .53, p *<* .001). Lastly, cerebellar network only had correlation to motor concept (r = .35, p *<* .001). The right frontoparietal and medial visual networks had no significant correlations to any of the concept maps.

**Figure 2:**
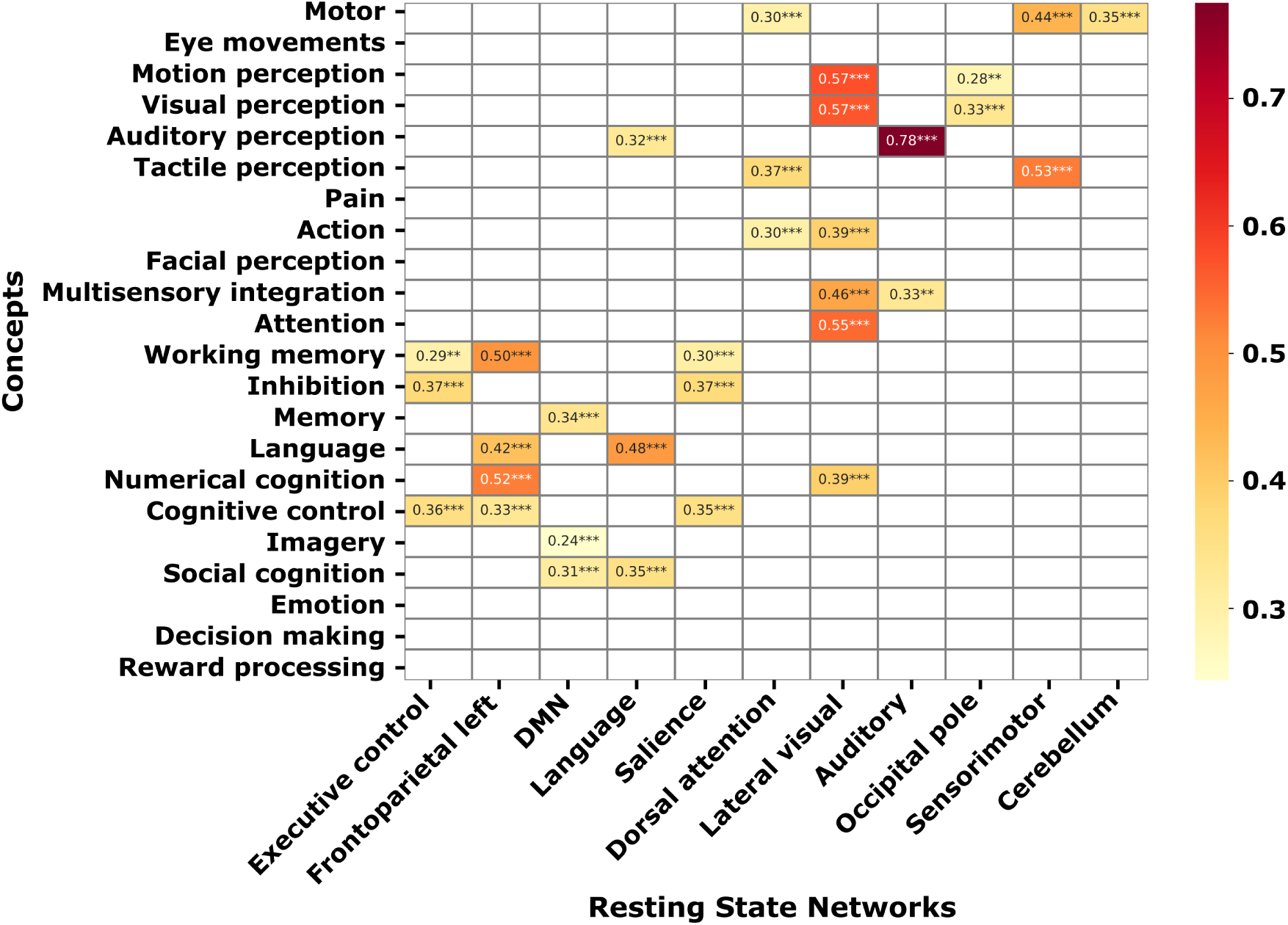
Correspondence between classical Resting-State Networks and Concept Maps. Correlation heatmap between Neurosynth-derived concept maps (y-axis) and HCP ICA-based RSN maps (x-axis). Concepts are ordered from lower-order, unimodal functions at the top, to higher-order cognitive functions at the bottom. RSN maps are arranged from higher-order networks (left) to lower-order, sensory/motor networks (right). Color intensity represents correlation strength, with warmer colors indicating stronger positive correlations. All correlations are significant when compared to a brain-surrogate null distribution, with * = p *<* .05, ** = p *<* .01, *** = p *<* .001.

### 3.2 Concept to disconnection representation (CDR)

For each patient, we computed CDRs between every concept map and the corresponding lesion-correlated and lesion-anticorrelated connectivity maps, generating an individual table of values. Fig 3 shows a representative example. The accompanying plots list all concepts and annotate only those CDRs that remain significant after Bonferroni correction (p *<* 0.05). All CDRs were normalized to the patient-specific maximum within each dysconnectivity class, and this peak value is the one reported on the plot. To quantify cognitive concepts most often affected by stroke-related network disruption, we counted each concept’s occurrence among significant CDRs for the two dysconnectivity classes. In lesion-correlated maps, the most common associations were motor (189 occurrences, 65.2%), pain (115, 63.1%), and tactile perception (170, 58.6%) (Fig 4a). In lesion-anticorrelated maps, social cognition (171, 59.0%), memory (144, 49.7%), and emotion (100, 34.5%) dominated (Fig 4b).

**Figure 3:**
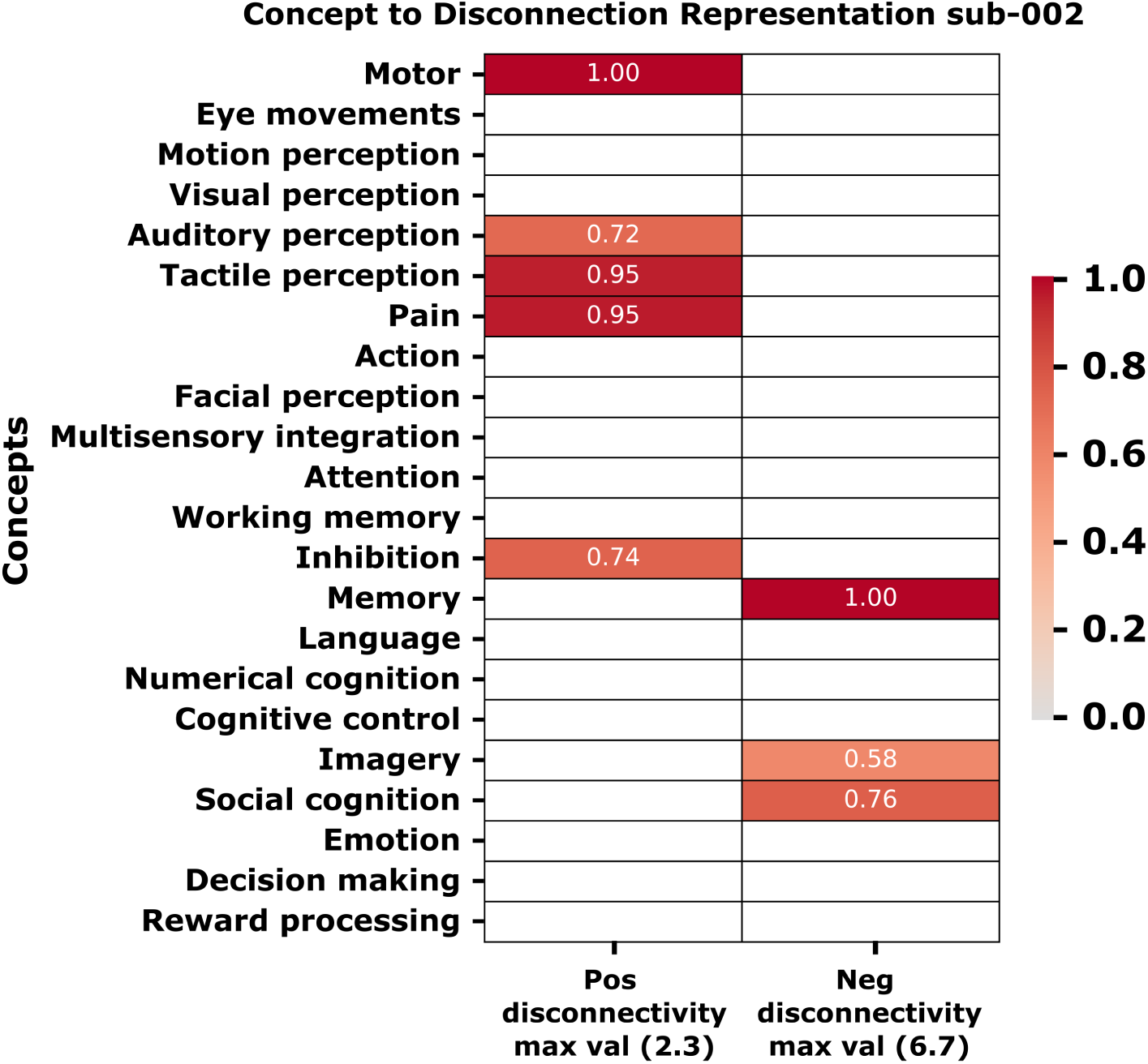
Concept to disconnection representation (CDR) report table example. All the concepts are ordered from lower-order, unimodal functions at the top to higher-order cognitive functions at the bottom. There are two columns in the table with the CDR values related to the positive dysconnectivity and the negative. CDR values are shown only if they are significant. Those values are normalized by dividing themby the maximum, and the maximum is also reported in the x-axis (in this example the maximums were 2.3 (motor) for positive and 6.7 (memory) for negative dysconnectivities).

**Figure 4:**
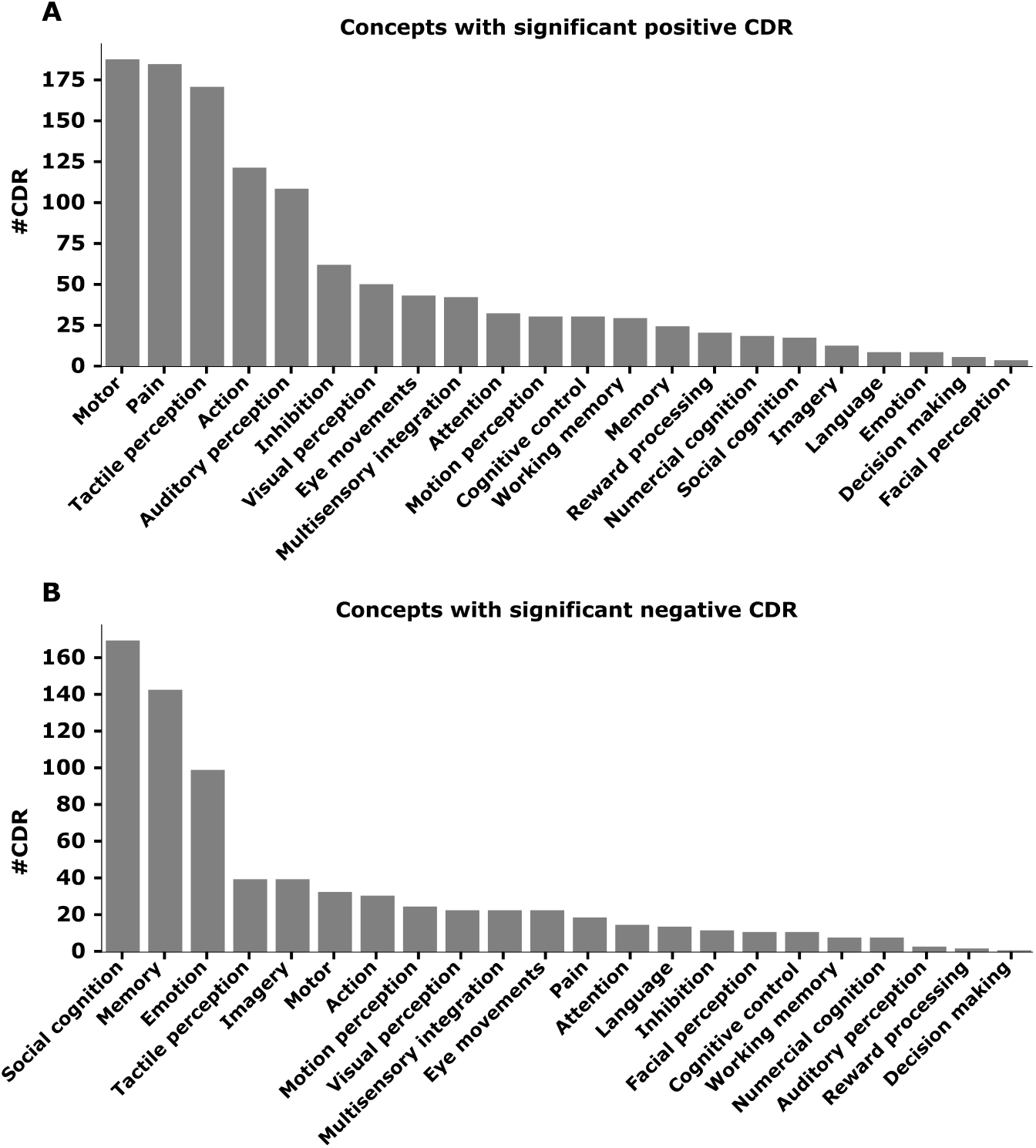
Concept alteration frequency across stroke patients. Each bar represents the number of patients with each concept significantly altered. A: concepts altered in relation to the positive lesion-dysconnectivity. B: concepts altered related to the negative lesion-dysconnectivity.

### 3.3 Prediction validation

Figure 5 shows NIHSS sub-item scores across patients whose CDR values were significant and those whose values were not, separately for the motor and sensory concepts. This analysis was restricted to patients for whom the relevant NIHSS sub-item scores were available. We found 76 patients exhibiting a significant motor CDR (and of those, only 68 had discharge NIHSS sub-items), whereas 37 did not have them neither at admission or discharge. The significant-CDR group had higher NIHSS-motor scores at admission (T = 4.57, p *<* 0.001) and discharge (T = 3.66, p *<* 0.001), indicating more severe motor deficits in those patients predicted by our tool. When network dysconnectivity information is discarded and predictions relied solely on lesion location, the model’s predictive value was poor, as we only found 15 patients whose lesions intersected the motor-concept mask, and their NIHSS motor scores were statistically indistinguishable from those without overlap, confirming the superior sensitivity of network-disconnection metrics over lesion-based approaches.

**Figure 5:**
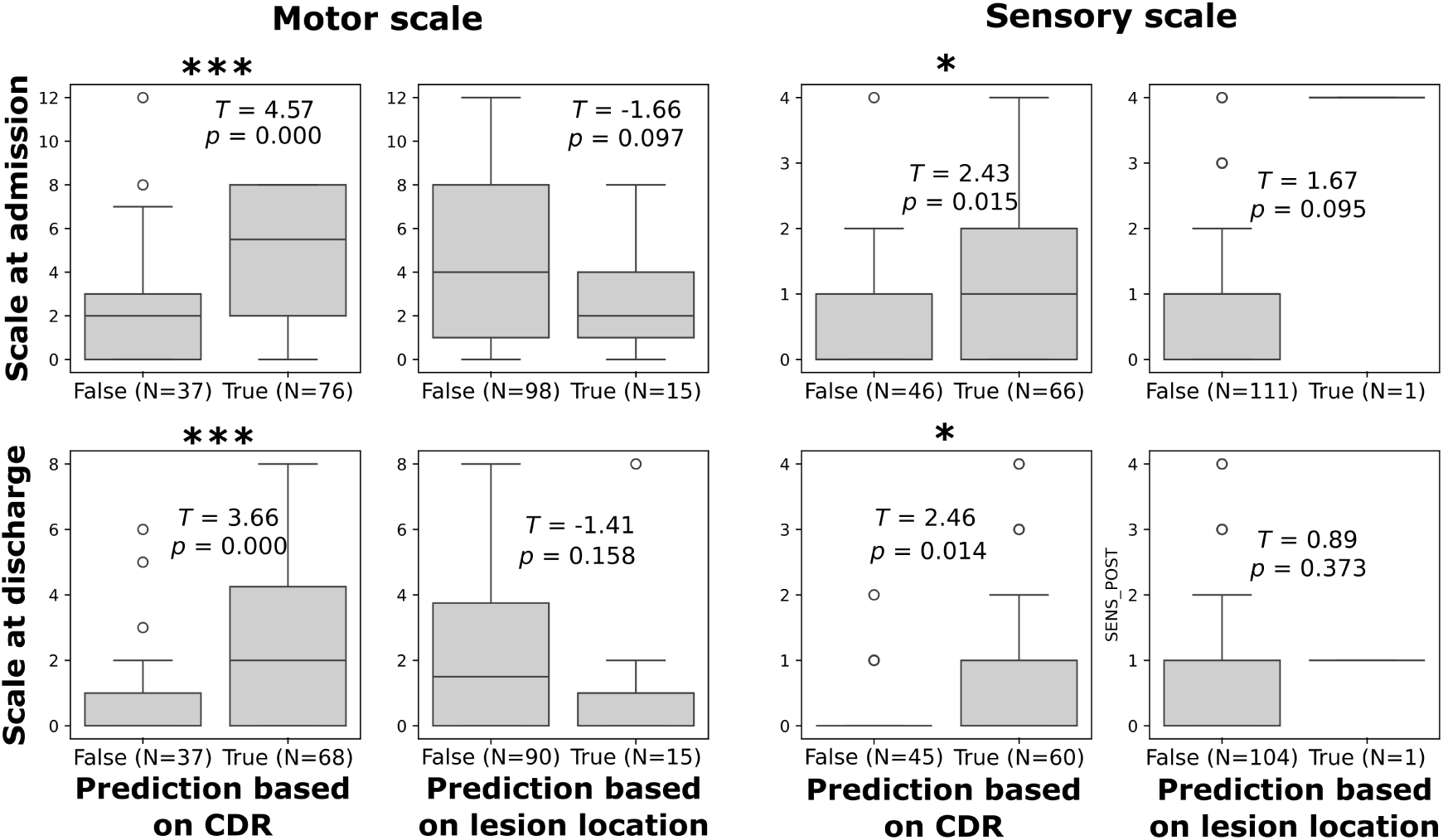
Validation of the COGNET-STROKE tool for motor and sensory scales. The scales were extracted from the NIHSS assessment at admission and discharge from the hospital. For both scales, boxplots with the scores are represented for those patients with significant CDR (after null-concept mask surrogates) and without it (left columns). P-values and the value of the statistic are also reported in the figure for every comparison. Furthermore, we represented the results obtained for the lesion significant overlap (with a similar mask surrogate strategy) as a comparison analysis (right columns). Therefore, for the sensory scale at admission and discharge only one subject had significant lesion overlap due to the limited specificity of the approach.

Similar findings emerged for the sensory NIHSS sub-item. At admission, 66 patients exhibited significant sensory CDRs, whereas 46 did not; the CDR-positive cohort demonstrated significantly greater sensory impairment (T = 2.43, p = 0.015). At discharge, 60 patients retained significant sensory CDRs and 45 did not, reproducing the admission pattern (T = 2.46, p = 0.014). A control analysis based solely on lesion overlap identified a single patient with concordant sensory involvement, confirming the negligible sensitivity of lesion-location based metrics.

## 4 Discussion

This study introduces COGNET-STROKE, a novel tool that uses a large database to facilitate datadriven interpretations of dysconnectivity patterns in stroke patients, and predict cognitive deficits associatted to network alterations in these patients. The tool operates by analyzing the relationship between meta-analytically derived concept maps from Neurosynth and functional dysconnectivity maps generated through lesion network mapping. Its foundational validity was evaluated through a two-step strategy: (i) spatial correlations were quantified between each concept map and canonical resting-state networks (RSNs) from the Human Connectome Project (networks previously shown to mirror task-evoked activation patterns (Smith et al., 2009)) and (ii) stratifying patients into groups predicted by COGNET-STROKE to exhibit motor or sensory impairment versus no impairment yielded significant between-group differences in the corresponding domain-specific behavioral scores.

For the first validation step, the RSN maps were obtained through performing an ICA on the same HCP dataset used to create the LNM maps. This allowed us to verify that concept maps corresponded to established large-scale brain networks, ensuring their interpretability and relevance for cognitive processes. The executive control network, often referred to as the cognitive control network or cognitive-executive network showed correlations to reward processing, decision making, emotion and social cognition. This network is known to activate during cognitive functions such as attention and working memory (Corbetta & Shulman, 2002; Mulders et al., 2015), and shows strong task-related activation, which explains the correlation to cognitive control, working memory and inhibition.

Working memory, numerical cognition, language, and cognitive control were all correlated with the frontoparietal network. This network is involved in sustained attention, complex problem solving and working memory (Menon, 2011), which validates the correlations to working memory and cognitive control. The correlation to language might be explained by the frequent co-activation of parts of the language end frontoparietal network when performing language related tasks (Fedorenko & Thompson-Schill, 2014). Furthermore, the frontoparietal network also often co-activates during numerical cognition tasks (Moeller et al., 2015), explaining the correlation to that concept.

The DMN is known to activate during passive rest and mind wandering. This usually involves remembering the past, thinking about others or oneself, and envisioning the future (Andrews-Hanna, 2012; Buckner et al., 2008; Xu et al., 2016). Furthermore, the role of the DMN has been associated to different social processes, such as theory of mind and moral reasoning (Andrews-Hanna, 2012). Thus, the correlations to imagery, social cognition and memory are valid upon theoretical review.

The correlation of the language network with the language concept might be explained directly with the name of both, network and concept. Social cognition and language has been reported with overlapped neural correlates (Zhang et al., 2023).

The salience network showed correlations to working memory, cognitive control and inhibition concepts. This could be related to the fact that the salience network has been described as the modulator between the dorsal and ventral attention networks, enabling dynamic control of attention in relation to top-down goals and bottom-up sensory stimulation (Vossel et al., 2014).

In relation to the correlations with the dorsal attention network, its role in top-down orientation and maintenance of visuospatial attention, motor and action could be due to the function of directing visual attention (Corbetta et al., 2000; Kincade et al., 2005).

The lateral visual network showed strong positive correlations to motion perception, visual perception, and multisensory integration. All of these concepts have strong implication in processes known to be situated in the visual cortex, embedded in the visual network (Seitzman et al., 2019; Thomas Yeo et al., 2011). Furthermore, research shows that functional connectivity of the visual network is closely tied to the fusiform face area in the brain, responsible for face processing (Haxby et al., 2000; M. Kim et al., 2006; Maher et al., 2019). The presence of a correlation to numerical cognition is no surprise either, since research by (de Hevia et al., 2008) shows that the visual cortex of the brain is connected to, and used by, visuo-spatial processes when performing numerical abilities. Moreover, the visual network is also known to have ties to action (Reetz et al., 2012) and attention processes (Parks & Madden, 2013; Ptak, 2012).

For the auditory network, the correlations with auditory perception and multisensory integration concepts can be explained directly with the name of the network and concepts. The occipital pole network was associated with visual functions, including visuospatial processing and object recognition. The sensorimotor network (Smith et al., 2009), including the supplementary motor area, sensorimotor cortex and secondary somatosensory cortex is, as expected, correlated with motor and tactile perception concepts. Lastly, the cerebellum network had an association with the motor concept, which is also expected.

To investigate lesion-induced functional disruption, each patient’s LNM dysconnectivity profile was partitioned into lesion-correlated and lesion-anticorrelated components, and each component was correlated with 22 meta-analytic cognitive concept maps. We identified pain, motor, and tactile perception as the most significantly correlated concepts, underlying the importance of sensorimotor and pain-processing symptoms in the context of stroke induced brain lesions. These findings are consistent with previous work by (Carey et al., 2016) showing that one in two stroke survivors lose their sense of touch. (Harrison & Field, 2015) also showed that chronic pain syndromes were found in up to half of stroke patients (Naess et al., 2012), of which 70% experience pain on a daily basis (Klit et al., 2011).

Our analysis of lesion-anticorrelated concepts provided critical insights into brain regions that exhibit decreased activity in relation to the lesioned area. Unlike the correlated connectivity results, anticorrelated connectivity was more related to social cognition, memory and emotion. These findings could be related to Post-Stroke Cognitive Impairment which can occur in up to 60% of stroke survivors in the first year after stroke (El Husseini et al., 2023), leading memory problems and depression.

The individualized connectivity–concept profiles generated by COGNET-STROKE advance towards precision rehabilitation. Our tool delineates, for each patient, the specific cognitive domains and network nodes most perturbed by the lesion-dysconnectivity, providing clinicians novel insights to engineer targeted interventions. This protocol operationalizes the principles of personalized medicine, grounded in the clinically meaningful inter-individual heterogeneity of neural organization and cognition (Goetz & Schork, 2018), and translates them into data-driven, therapeutical intervention strategies.

We have validated COGNET-STROKE in two common neurological post-stroke symptoms: Motor and Sensory deficits. Both clinical scores have been extracted from the NIHSS assessment at admission in the hospital and at discharge, around 40 days after stroke. During the admission period, the patients received complete rehabilitation. Both scores were significantly lower at the two time-points in patients predicted to have motor or tactile perception impairment. To further validate the tool, we also tested if our results were better than a prediction based only on the location of the lesions, getting higher differences with our network-disconnection approach.

Other existing tools pursue aims analogous to COGNET-STROKE. The Lesion Quantification Toolkit (Griffis et al., 2021) makes use of lesion masks to compute four complementary indices—tract-wise dysconnectivity load for 70 white-matter bundles, voxel-wise dysconnectivity maps, network-level load within the 400-region Schaefer parcellation, and “indirect” disconnection via shortest path analysis (indirect meaning connectivity estimated on a normative cohort distinct from the patient sample). The tool provides a comprehensive damage/disconnection report whose parcels correlate with behavioral scores after Bonferroni correction. (Talozzi et al., 2023) provides lesion-derived dysconnectivity from five independent cohorts (N = 1133, 119, 26, 190, 193) into a two-dimensional UMAP space and trains predictive models for 86 neuropsychological tests at one-year follow-up, generating patient-specific cognitive profiles grounded in the HCP-7 T normative connectome. (Thiebaut de Schotten et al., 2020) applies an identical UMAP embedding to 1,333 lesions to construct an atlas of white-matter circuits functionally annotated with Neurosynth terms; lesion-behavior associations are inferred by projecting neuropsychological scores onto this atlas, referencing the HCP-7 T normative dataset. Advanced Lesion Symptom Mapping Analyses Implemented as BCBtoolkit (Foulon et al., 2018) integrates white-matter dysconnectivity and indirect functional connectivity extracted from lesion masks to relate disconnection patterns to category and letter fluency in 37 patients, showing significant group differences and employing a normative sample of 10 healthy controls to estimate indirect connectivity.

A first methodological limitation is related to the LNM framework, which is based on normative connectomes, allowing only a generalized characterization of lesion-induced network dysconnectivity and does not capture inter-individual variability in the absence of patient-specific functional data. However, the major limitation of our study is sample composition, where all patients were drawn from the rehabilitation and physical medicine service at Gorliz Hospital and were characterized by marked, multisystem functional impairment. Consequently, we lack patients with relatively focal cognitive syndromes (e.g., isolated aphasia, selective attentional deficits), constraining our ability to validate the tool across discrete cognitive domains beyond the motor and sensory impairments that predominate in this cohort. Moreover, the absence of domain-specific neuropsychological assessments makes it challenging to assess whether lesion-based concept scores truly reflect behavioral deficits. However, the motor and sensory validation suggests that the approach may have some degree of reliability in capturing cognitive impairment. On the other hand, our results regarding the validation of the sensory scale corresponded to the concept of tactile perception, although we know that our scale encompasses the sum of tactile perception and other sensory neglects. Our validation is partially limited by this ambiguity. Regarding the predictive capability of our tool, our results are based on the association between disconnection patterns and clinical scales at the beginning and at the end of rehabilitation. Beyond this association, prognostic studies are needed to evaluate the efficiency of the prediction. Future studies should also include a more heterogenous sample of patients, together with a more comprehensive battery of cognitive tests to strengthen validation efforts, possibly also using longitudinal data to explore how these variables evolve over time.

## 5 Conclusion

This study presents COGNET-STROKE, a novel tool for understanding stroke-related cognitive deficits by integrating LNM with meta-analytic functional decoding. By leveraging Neurosynth-derived cognitive concept maps and LNM-derived dysconnectivity maps, we identified cognitive processes most affected by lesion-induced network disruptions. Our framework identifies cognitive domains associated with lesion-induced functional dysconnectivity and shows initial behavioral relevance for motor and sensory impairments in this cohort.

## Data Availability

MRI images used from the 1000 healthy subjects are available under registration at https://www.humanconnectome.org/.
Clinical data is available upon reasonable request.

## Data and Code Availability

MRI images used from the 1000 healthy subjects are available under registration at. https://www.humanconnectome.org/. Clinical data is available upon reasonable request.

All the code used in the article is openly available on our lab GitHub Repository: https://github.com/ compneurobilbao, in particular:

- For registering the CT images to the MNI152 standard space, we applied *CTLesion2MNI152* (https://github.com/compneurobilbao/CTLesion2MNI152).
- For generating the dysconnectivity maps we applied *lnm functional*, available on https://github.com/compneurobilbao/lnm.
- The COGNET-STROKE tool and all the analyses and code involved in the methods and results of this manuscript is available on https://github.com/compneurobilbao/COGNET-STROKE.

## Author Contributions

A.J.M. and S.B. contributed equaly to conceptualization, data curation, formal analysis, methodology, software, and visualization. I.T.E. contributed to data curation, formal analysis, methodology, and software. I.E. contributed to resources, supervision, and validation. A.C.Z. contributed to resources, supervision, and validation. G.D.S. contributed to conceptualization, data curation, investigation, and resources. M.F. contributed to conceptualization, resources, supervision, and validation. P.I.T. contributed to conceptualization, resources, investigation, supervision, and validation. R.T. contributed to methodology, software, and supervision. I.D. contributed to formal analysis, methodology, software, and supervision. A.E. contributed to conceptualization, formal analysis, funding acquisition, investigation, methodology, project administration, software, supervision. J.M.C. contributed to conceptualization, formal analysis, funding acquisition, investigation, methodology, project administration, software, supervision. All authors contributed to writing the original draft.

## Acknowledgements

JMC acknowledges financial support from the Spanish Ministry of Health (PI22/01118) and Basque Ministry of Health (2023111002 & 2022111031). JMC and AE are both funded by the Spanish Ministry of Science (grant PID2023-148008OB-I00). ID was supported by the Spanish Ministry of Science with the grant PID2023-150633OA-I00. AE and ID are both funded by the Spanish Ministry of Science and Innovation grants RYC2021-032390-I and RYC2022-035429-I, respectively. JMC, AE, and ID are funded by Ikerbasque: The Basque Foundation for Science. MMF acknowledges financial support from Carlos III Institute of Health (grant RICORS-Ictus RD24/0009/0004).

